# Screening for Right Ventricular Dysfunction in the Emergency Department Using a Smartphone ECG Analysis Application: An External Validation Study with Acute Pulmonary Embolism Patients

**DOI:** 10.1101/2024.05.08.24307018

**Authors:** Yoo Jin Choi, Min Ji Park, Youngjin Cho, Joonghee Kim, Eunkyoung Lee, Dahyeon Son, Seo-Yoon Kim, Moon Seung Soh

**Affiliations:** Department of Emergency Medicine, Ajou University School of Medicine, Suwon-si 16499, Republic of Korea; Department of Cardiology, Ajou University School of Medicine, Suwon-si 16499, Republic of Korea; Cardiovascular Center, Department of Internal Medicine, Seoul National University Bundang Hospital, 166 Gumi-ro, Bundang-gu, Seongnam-si, Gyeonggi-do, 13620, Republic of Korea; Department of Emergency Medicine, Seoul National University Bundang Hospital, 166 Gumi-ro, Bundang-gu, Seongnam-si, Gyeonggi-do, 13620, Republic of Korea; ARPI Inc., Room 12 Startup Incubation Center, 172, Dolma-ro, Bundang-gu, Seongnam-si, Gyeonggi-do, 13605, Republic of Korea

**Keywords:** RV dysfunction, Pulmonary embolism, Digital biomarkers, ECG analysis application, Emergency department

## Abstract

**Background:** Acute pulmonary embolism (PE) is a critical condition where timely and accurate assessment of right ventricular (RV) dysfunction is important for patient management. Given the limited availability of echocardiography in emergency departments (EDs), an artificial intelligence (AI) application that can identify RV dysfunction from electrocardiograms (ECG) could improve the treatment of acute PE.

**Methods:** This retrospective study analyzed adult acute PE patients in an ED from January 2021 to December 2023. We evaluated a smartphone application which analyzes printed ECG to generate digital biomarkers for various conditions including RV dysfunction (QCG-RVDys). The biomarker’s performance was compared with that of cardiologists and emergency physicians.

**Results:** Among 116 included patients, 35 (30.2%) were diagnosed with RV dysfunction. The QCG-RVDys score demonstrated significant effectiveness in identifying RV dysfunction, with a receiver operating characteristic - area under the curve(AUC) of 0.895 (95% CI, 0.829-0.960), surpassing traditional biomarkers such as Troponin I (AUC: 0.692, 95% CI: 0.536-0.847) and ProBNP (AUC: 0.655, 95% CI: 0.532-0.778). Binarized based on the Youden Index, QCG-RVDys achieved an AUC of 0.845 (95% CI: 0.778-0.911), with sensitivity, specificity, positive predictive value (PPV), and negative predictive value (NPV) of 91.2% (95% CI: 82.4-100%), 77.8% (95% CI: 69.1-86.4%), 63.3% (95% CI: 54.4-73.9%), and 95.5% (95% CI: 90.8-100%), respectively, significantly outperforming all the expert clinicians with their AUCs ranging from 0.628 to 0.683.

**Conclusion:** The application demonstrates promise in rapidly assessing RV dysfunction in acute PE patients. Its high NPV could streamline patient management, potentially reducing the reliance on echocardiography in emergency settings.

## Introduction

Acute pulmonary thromboembolism (PE) is a serious emergency condition that can be life-threatening. Screening right ventricular (RV) dysfunction is crucial in the management of acute PE[1], as it significantly impacts patient outcomes. The RV’s response to increased afterload can result in myocardial ischemia and heart failure, increasing the risk of hemodynamic collapse and mortality[2]. Therefore, timely and accurate detection of RV dysfunction is vital for risk stratification and guiding treatment in acute PE.

Echocardiography is the standard tool for evaluating RV function, providing detailed hemodynamic information [3–5]. However, the fast-paced environment of the emergency department (ED) requires faster and simpler diagnostic methods, and in that regard, echocardiography may be limited in its use in the ED due to its reliance on skilled operators and the availability of equipment.

One potential solution to address this issue is to utilize electrocardiograms (ECGs). Recognizing RV strain patterns such as new right-axis deviation, the S1Q3T3 pattern, or ST-segment depressions with T-wave inversions in leads V1 to V3 and leads II, III, and aVF from ECGs, can facilitate the evaluation of RV dysfunction[6,7]. However, these methods have limitations in accuracy, reducing their utility.

The integration of digital technology into medicine, especially the use of artificial intelligence (AI) in acute care, offers new possibilities. AI solutions that analyze ECG data represent a shift toward more accessible and rapid cardiac assessments[8,9]. However, these applications typically require raw digital ECG data, which is impractical in real-world clinical settings where only printed ECG data is available.

To bridge this gap, we developed ECG Buddy^TM^, a mobile application that generates ten digital biomarkers by analyzing images of printed ECGs. Previous studies have suggested this tool’s utility in various emergency situations including suspected myocardial infarctions and severe hyperkalemia [10–13]. This study aims to evaluate the application’s capability in identifying RV dysfunction in ED patients with acute PE. Additionally, we will compare its performance to those of expert clinicians.

## Methods

### 2.1. **Study design and data collection**

This retrospective study analyzed adult patients (≥18 years) with acute PE presenting at the ED of a tertiary hospital from January 2021 to December 2023. Patients were excluded if they did not have an ECG within 72 hours post-ED arrival or an echocardiogram from 72 hours before to 72 hours after the ECG. Two board-certified emergency physicians manually reviewed electronic medical records (EMR) to determine eligibility and gather information on demographics, PE risk factors, vital signs, lab results, and heart rhythm. The ECG data was collected from the EMR, by manually cropping the waveform areas of each ECG reports to remove any identifying information. The institutional review board approved the study (IRB No.: AJOUIRB-DB-2024-177), waiving informed consent due to its retrospective design.

### 2.2 Assessment of RV function

The presence of RV dysfunction was determined by qualitative review of echocardiographic reports. The RV dysfunction was defined as a two-dimensional fractional area change (FAC) of less than 35% according to the literature. FAC was calculated as (end-diastolic area-end-systolic area)/end-diastolic area x 100 in the RV-focused view on echocardiogram [5]. In addition, measurements of echocardiographic right ventricular systolic pressure (RVSP), an indicator of pulmonary hypertension, were collected and categorized into four groups: RVSP I (less than 35 mmHg), RVSP II (35-49 mmHg), RVSP III (50-64 mmHg), and RVSP IV (greater than 64 mmHg). RVSP was determined from peak tricuspid regurgitant jet velocity (TR Vpeak), using the simplified Bernoulli equation and combining this value with an estimate of the right atrium (RA) pressure estimated from inferior vena cava diameter and respiratory changes in the subcostal view [RVSP=4(TR Vpeak)^2^+RA pressure].

### 2.3. ECG analysis by AI application

An AI smartphone application, named “ECG Buddy”, was used to analyze ECG images. The application, approved by the Korean MFDS and freely available for download in Korean appstores, can analyze 12-lead ECGs by taking picture of ECG images to produce 10 digital biomarkers, (Quantitatve ECG[QCG®] scores, ranging from 0 to 100) for various emergencies and cardiac dysfunctions.[14] We analyzed the ECG capture images by first displaying them on desktop monitor and taking picture of the ECG images using the application. We recorded digital biomarkers for RV dysfunction (QCG-RVDys) and pulmonary hypertension (QCG-PHTN) for each ECG for evaluation.

### 2.4 Expert Analysis of ECGs

To establish a benchmark for the AI biomarkers, expert evaluations of the same ECGs were obtained from a group of two cardiologists and three emergency physicians, all blinded to the patients’ clinical information. The experts, all board-certified physicians with at least 8 years of clinical experiences, asked to review each ECG images freely without time limit to determine whether the ECG exhibited RV dysfunction using primarily S1Q3T3 pattern[15,16].

### 2.5. Statistical Analysis

The primary metric used to assess the performance of the biomarkers was receiver operating characteristic - area under the curve (ROC-AUC). We compared the ROC-AUC of QCG-RVDys for identifying RV dysfunction to those for troponin I and proBNP using original measurements in continuous scale. For comparison with experts, we binarized the biomarker using the threshold that maximized its Youden index (Binarized QCG-RVDys) as the experts were asked to give their opinion in binary format (yes or no). Sensitivity, specificity, positive predictive value (PPV), and negative predictive value (NPV) of the biomarker were calculated. The ROC-AUC of QCG-PHTN in identifying moderate pulmonary hypertension, as determined by RVSP 50mmHg or more, was calculated too, however, the performance of the biomarker was not compared to other measurements as it is not the main study topic of the study and there is no popular method for identifying the condition clinically. All data analysis was conducted using R software, version 4.1.0.

## Results

From January 2021 to December 2023, 131 patients with acute PE were admitted to the emergency room. After exclusion of fifteen patients lacking ECG or echocardiography measurements meeting the eligibility criteria, a total of 116 patients were included in the study (Fig. 1). Within the patient population, 35 patients were assessed as having RV dysfunction (RVD group, 30.2%) and 81 patients were assessed as not having RV dysfunction (No RVD group, 69.8%).

**Fig. 1.**
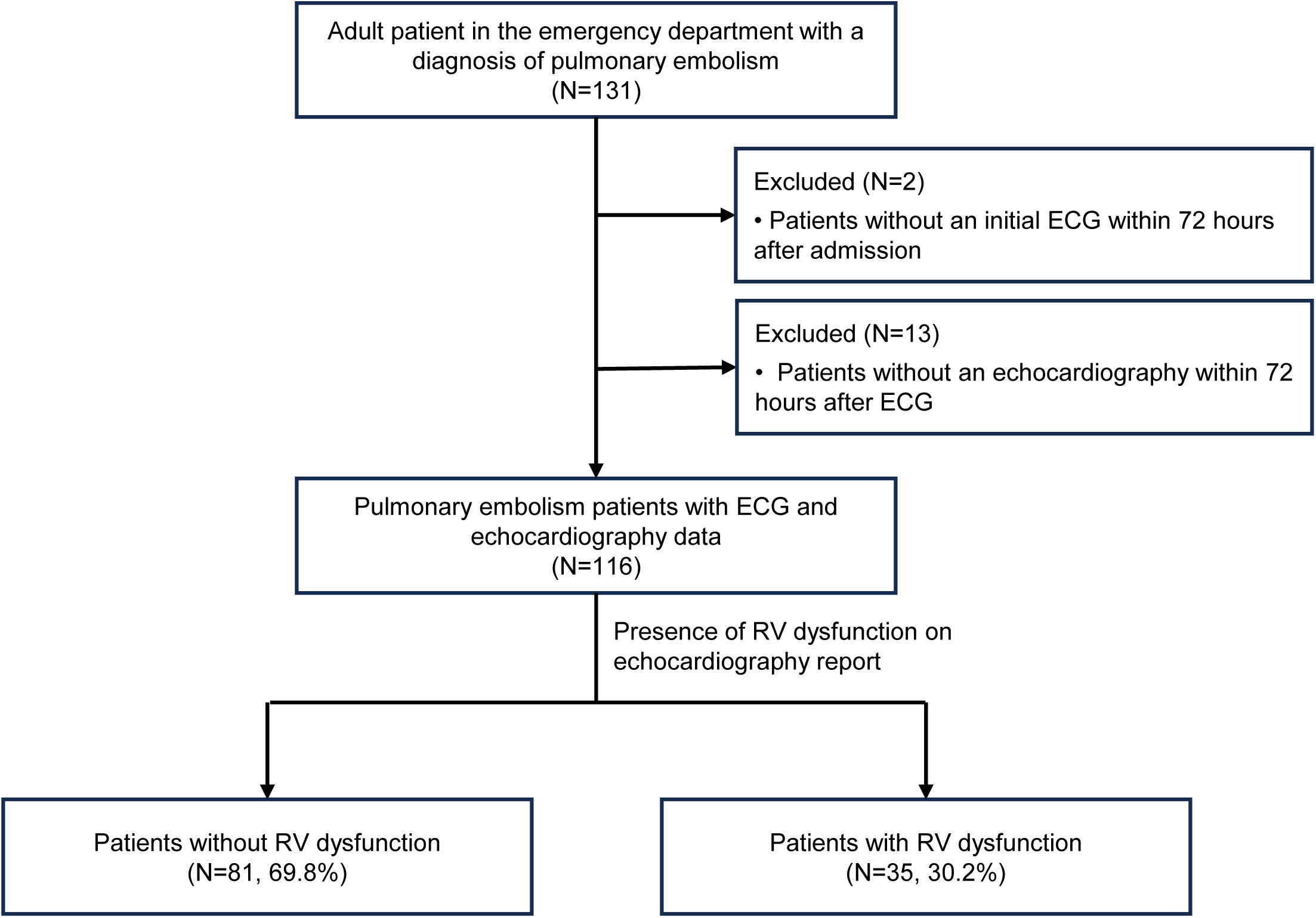
Patient selection flowchart

There were no statistically significant differences between the two groups in terms of previous risk factors, except for the number of patients with a history of deep vein thrombosis (No RVD: 0, RVD: 3, p=0.042) (Table 1). Initial vital signs measured in the ED showed no significant differences. However, in blood tests, aspartate transaminase (AST; No RVD: 22.5U/L vs. RVD: 32.0U/L, P= 0.003), alanine transaminase (ALT; No RVD: 19.0U/L vs. RVD: 30.0U/L, P= 0.017), Troponin I (No RVD: 0.052 ㎍/mL vs. RVD: 0.176 ㎍/mL, P= 0.034), and proBNP (No RVD: 482.0pg/mL vs. RVD: 1366.0pg/mL, P=0.024) were significantly higher in the RVD group. In addition, RVSP group distribution was also significantly different (P<0.001), and there was a trend for echocardiography to be performed earlier in the RVD group (ED arrival to echocardiography, No RVD: 22.1 hours vs. RVD: 16.1 hours, P= 0.036).

**Table 1.** Patient characteristics

|  |  | Presence of RV dysfunction |  |  |
| --- | --- | --- | --- | --- |
|  |  | No (N=81) | Yes (N=35) |  |
| Demographics | Age, years, median (IQR) | 72.0 (50.0-81.0) | 67.0 (45.5-77.0) | 0.388 |
|  | Sex, male, N (%) | 35 (43.2%) | 14 (40.0%) | 0.907 |
|  | Weight, kilograms, median (IQR) | 61.3 (50.2-73.8) | 55.8 (53.0-74.0) | 0.946 |
|  | Height, centimeters, median (IQR) | 160.0 (155.0-170.0) | 162.0 (157.0-168.0) | 0.781 |
| Risk Factors of PTE | Diabetes Mellitus, N (%) | 18 (22.2%) | 5 (14.3%) | 0.465 |
|  | Hypertension, N (%) | 37 (45.7%) | 12 (34.3%) | 0.350 |
|  | Coronary artery occlusive disease, N (%) | 7 (8.6%) | 3 (8.6%) | 1.000 |
|  | Cerebrovascular disease, N (%) | 10 (12.3%) | 1 (2.9%) | 0.209 |
|  | Current Smoker, N (%) | 5 (6.2%) | 3 (8.6%) | 0.945 |
|  | Prolonged immobility (>1 week), N (%) | 18 (22.2%) | 8 (22.9%) | 1.000 |
|  | Recent trauma or surgery (within 3 months), N (%) | 16 (19.8%) | 13 (37.1%) | 0.080 |
|  | Active malignancy, N (%) | 18 (22.2%) | 3 (8.6%) | 0.136 |
|  | Infectious disease (within 3 months), N (%) | 19 (23.5%) | 3 (8.6%) | 0.105 |
|  | Hormone treatment, N (%) | 2 (2.5%) | 1 (2.9%) | 1.000 |
|  | History of pulmonary thromboembolism, N (%) | 10 (12.3%) | 4 (11.4%) | 1.000 |
|  | History of deep vein thrombosis, N (%) | 0 (0.0%) | 3 (8.6%) | 0.042 |
| Vital Signs | Systolic blood Pressure, mean (SD) | 130.0 (26.1) | 129.4 (26.6) | 0.915 |
|  | Diastolic blood pressure, mean (SD) | 82.2 (17.7) | 81.0 (15.1) | 0.734 |
|  | Pulse rate, median (IQR) | 99.0 (81.5-115.5) | 101.0 (90.0-117.0) | 0.150 |
|  | Respiratory rate, median (IQR) | 20.0 (18.0-22.0) | 21.0 (20.0-24.0) | 0.092 |
| Laboratory Measurements | White blood cell, 10<sup>9</sup>/L, median (IQR) | 9.2 (7.4-13.1) | 10.0 (7.3-13.5) | 0.740 |
|  | Hemoglobin, g/dL, mean (SD) | 12.1 (2.4) | 12.4 (2.3) | 0.486 |
|  | Aspartate transaminase, U/L, median (IQR) | 22.5 (16.0-35.0) | 32.0 (25.0-56.0) | 0.003 |
|  | Alanine transaminase, U/L, median (IQR) | 19.0 (12.0-34.0) | 30.0 (18.0-60.5) | 0.017 |
|  | Blood urea nitrogen, mg/dL, median (IQR) | 14.9 (11.0-22.4) | 14.3 (11.5-19.6) | 0.551 |
|  | Creatinine, mg/dL, median (IQR) | 0.9 (0.7-1.1) | 0.8 (0.7-1.1) | 0.440 |
|  | Troponin I, µg/mL, median (IQR) | 0.052 (0.019-0.266) | 0.176 (0.075-0.723) | 0.034 |
|  | ProBNP, pg/mL, median (IQR) | 482.0 (140.0-2400.0) | 1366.0 (576.0-4733.0) | 0.024 |
|  | D-dimer, mg/L, median (IQR) | 3.6 (2.1-12.3) | 4.6 (2.8-10.7) | 0.599 |
|  | Lactate, mg/dL, median (IQR) | 1.0 (0.8-2.3) | 2.0 (1.1-2.3) | 0.097 |
| Heart rhythm (on ECG) |  |  |  | 0.117 |
|  | Sinus Rhythm, N (%) | 52 (64.2%) | 14 (41.2%) |  |
|  | Sinus Tachycardia, N (%) | 22 (27.2%) | 16 (47.1%) |  |
|  | Atrial Fibrillation, N (%) | 3 (3.7%) | 1 (2.9%) |  |
|  | Multifocal Atrial Tachycardia, N (%) | 2 (2.5%) | 0 (0.0%) |  |
|  | Sinus Arrhythmia, N (%) | 1 (1.2%) | 0 (0.0%) |  |
|  | Atrial Rhythm, N (%) | 0 (0.0%) | 1 (2.9%) |  |
|  | Wandering Atrial Rhythm, N (%) | 0 (0.0%) | 1 (2.9%) |  |
|  | Undetermined Rhythm, N (%) | 1 (1.2%) | 1 (2.9%) |  |
| Right Ventricular Systolic Pressure (RVSP) |  |  |  | <0.001 |
|  | RVSP I (<35 mmHg), N (%) | 61 (75.3%) | 4 (11.4%) |  |
|  | RVSP II (35-49 mmHg), N (%) | 15 (18.5%) | 12 (34.3%) |  |
|  | RVSP III (50-64 mmHg), N (%) | 5 (6.2%) | 10 (28.6%) |  |
|  | RVSP IV (>64 mmHg), N (%) | 0 (0.0%) | 9 (25.7%) |  |
| Time of the test | ED arrival to ECG, hours, median (IQR) | 1.0 (0.6-1.6) | 0.8 (0.6-1.3) | 0.388 |
|  | ED arrival to echocardiography, hours, median (IQR) | 22.1 (16.0-35.6) | 16.1 (4.8-27.5) | 0.036 |
|  | ECG to echocardiography, hours, median (IQR) | 20.4 (14.9-34.0) | 13.8 (4.3-26.1) | 0.045 |

RV, right ventricular; IQR, interquartile range; PTE, Pulmonary thromboembolism; SD, standard deviation; BNP, Blood natriuretic peptide; ECG, Electrocardiogram; ED, Emergency department

The QCG-RVDys scores were significantly different between the No RVD group (6.8 [2.5-22.5]) and the RVD group (78.7 [35.7-94.9]), with a p-value of <0.001 (Fig. 2, supplementary table 1). The QCG-PHTN scores also showed significant differences across the RVSP groups I, II, III, and IV, demonstrating a clear correlation with increasing RVSP (P<0.001, P-trend<0.001, supplementary Table 1 & supplementary Figure 1).

**Fig. 2.**
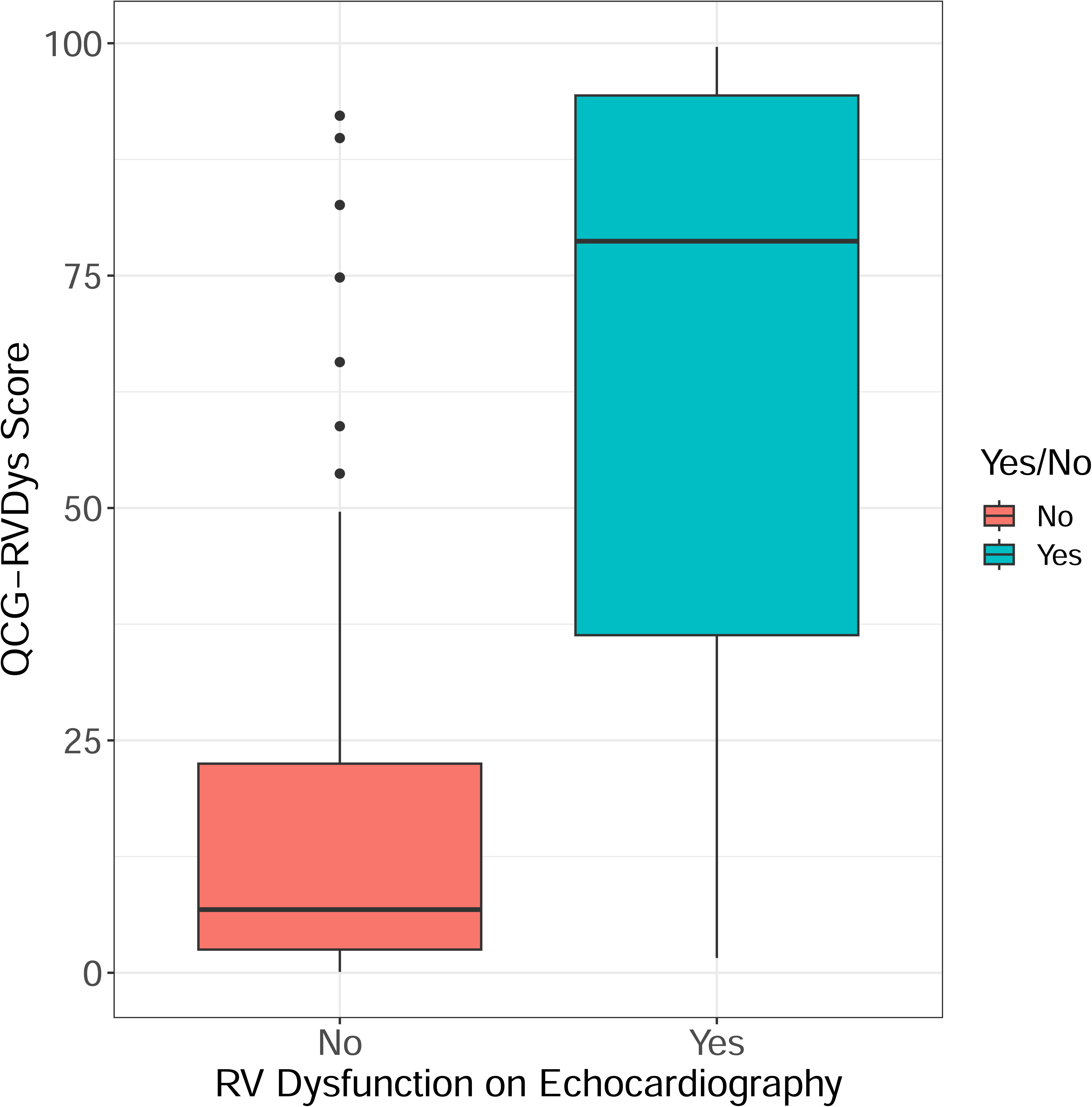
QCG score difference by RV dysfunction group

The AUC-ROC for QCG-RVDys in diagnosing RV dysfunction was 0.895 (0.829-0.960), significantly higher than that for Troponin I, 0.692 (0.536-0.847), and ProBNP, 0.655 (0.532-0.778) (p=0.046 and p=0.001, respectively, Fig. 3, table 2). When the QCG-RVDys was binarized at a threshold of 24.65, based on the Youden index, the AUC was 0.845 (0.778-0.911), with sensitivity, specificity, PPV, and NPV of 91.2% (82.4-100), 77.8% (69.1-86.4), 63.3% (54.4-73.9), and 95.5% (90.8-100), respectively. In comparison, the five experts showed AUCs ranging from 0.628 to 0.683, all statistically significantly lower than that of the binarized QCG-RVD.

**Fig. 3.**
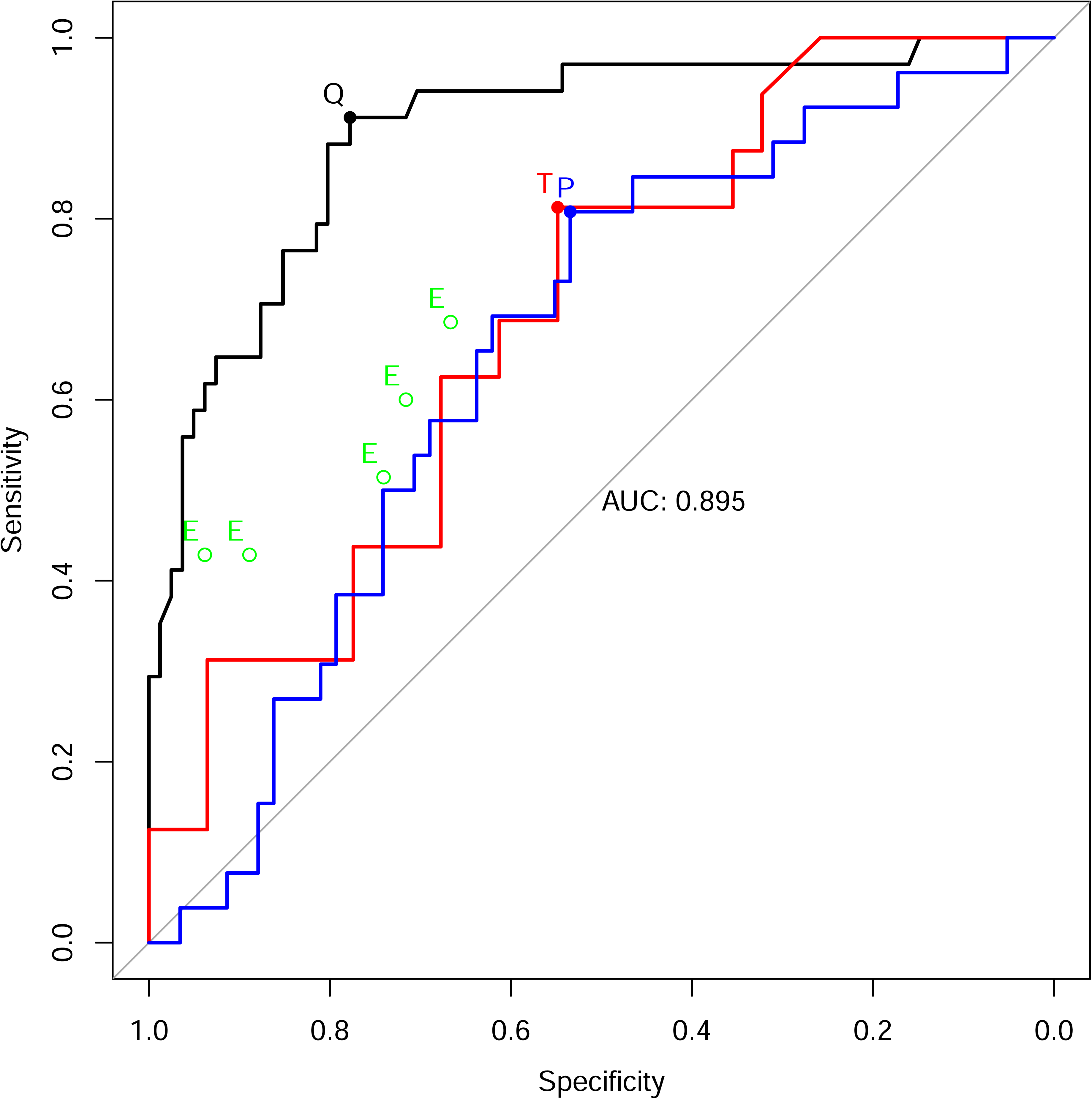
Performance of QCG-RVDys, Troponin I, ProBNP and clinical experts on identifying RV dysfunction from ECG (Black line, QCG-RVDys; Red line, Troponin I; Blue line, ProBNP; Green hollow dots, Experts; black red and blue dots indicate binarized conditions)

**Table 2.** Performance of QCG and other biomarkers for prediction of RV dysfunction

| Biomarker | AUC | P for difference | Sensitivity<br>(95% CI) | Specificity<br>(95% CI) | PPV<br>(95% CI) | NPV<br>(95% CI) | Threshold |
| --- | --- | --- | --- | --- | --- | --- | --- |
| QCG-RVDys (continuous scale) | 0.895 (0.829-0.960) | - | 91.2 (82.4-100) | 77.8 (69.1-86.4) | 63.3 (54.4-73.9) | 95.5 (90.8-100) | 24.65 |
| Troponin I | 0.692 (0.536-0.847) | 0.046 | 81.2 (62.5-100) | 54.8 (38.7-71) | 48.1 (37.5-61.1) | 85 (70.6-100) | 0.0685 µg/mL |
| ProBNP | 0.655 (0.532-0.778) | 0.001 | 80.8 (65.4-96.2) | 53.4 (39.7-65.5) | 43.8 (35.6-52.5) | 86.4 (75.8-96.3) | 534.5 pg/mL |

AUC, area under the curve; PPV, positive predictive value; CI, confidence interval; NPV, negative predictive value; QCG, quantitative electrocardiography; RVDys, right ventricular dysfunction; BNP, Blood natriuretic peptide

**Table 3.** QCG score difference by RV dysfunction and across RVSP groups

| QCG Biomarker | Group | Biomarker Measurements, median (IQR) | P |
| --- | --- | --- | --- |
| QCG-RVDys | RVD | 6.8 (2.5-22.5) | <0.001 (for difference) |
|  | No RVD | 78.7 (35.7-94.9) |  |
| QCG-PHTN | RVSP I (<35mmHg) | 8.1 (2.1-22.9) | <0.001 (for both difference and trend) |
|  | RVSP II (35-49mmHg) | 21.7 (13.4-38.3) |  |
|  | RVSP III (50-64mmHg) | 41.6 (31.2-65.1) |  |
|  | RVSP IV (>64mmHg) | 49.2 (19.9-85.4) |  |

QCG, quantitative electrocardiography; IQR, interquartile range; RVDys, right ventricular dysfunction; RVD, right ventricular dysfunction; PHTN, pulmonary hypertension; RVSP, right ventricular systolic pressure

The performance of QCG-PHTN in predicting elevated RVSP (RVSP groups III and IV; RVSP ≥ 50mmHg) showed a ROC-AUC of 0.820 (0.728-0.912) (Supplementary Fig. 2). At a threshold of 0.2590, the sensitivity, specificity, PPV, and NPV were 82.6% (65.2-95.7), 72.8% (64.1-81.5), 43.2% (34.7-54.1), and 94.4% (89.4-98.6), respectively.

## Discussion

This study showed that the digital ECG biomarker produced by a smartphone application can predict RV dysfunction with high accuracy in patients with acute PE using only printed ECGs without additional clinical information. We have also shown it can also predict pulmonary hypertension, too, expanding the utility of ECGs. This is the first study to assess critical cardiac functions in patients with acute PE using ECG AI, especially through a smartphone app, distinguishing it from similar research in the field.

Based on the results, smartphone-based ECG analysis software could be considered for use in treatment decisions for patients with acute PE in EDs. Assessing right heart function, especially RV dysfunction and pulmonary hypertension, is crucial for determining the treatment plan for acute PE, alongside evaluating hemodynamic stability. This can rapidly guide decisions regarding thrombolytic therapy or surgical embolectomy in patients with RV dysfunction and hemodynamic instability [2,17,18]. According to our study, QCG-RVDys has a high NPV of 95.5 (90.8 – 100%), suggesting it could be useful in excluding the possibility of RV dysfunction in patients with newly diagnosed acute PE for whom an immediate echocardiography is not feasible.

This utility can also be extended to the early diagnosis of massive PE. Patients with massive PE often present with nonspecific symptoms like dizziness, syncope, dyspnea, and chest pain, necessitating a broad differential diagnosis. In many EDs, when patients present with such symptoms, ECG is one of the first triage tests usually performed. Utilizing this, early detection of RV dysfunction, often associated with massive PE, could prioritize the suspicion of PE, allowing for early diagnostic imaging, such as contrast chest CT, and hastening diagnosis.[2] It has also been shown to be a good predictor of RV dysfunction compared to traditional markers that reflect cardiac burden, such as BNP and troponin, and is even more valuable given the time it takes to get results from blood tests.

Similarly, the evaluation of RV dysfunction could be expanded to other diseases, such as in acute respiratory distress syndrome and cor pulmonale, where assessment of RV strain is beneficial for establishing lung-protective ventilation strategies, balanced fluid therapy, selection of vasopressors (minimizing impact on pulmonary vascular resistance), pulmonary vasodilation (e.g., NO, sildenafil), and effective monitoring strategies (echocardiography or hemodynamic measurement)[19–23]. Likewise, early recognition of RV dysfunction in RV myocardial infarction through AI could aid safer decision-making regarding early fluid therapy and vasodilator use. However, the efficacy and safety of the application in these specific clinical scenarios have not been evaluated, indicating a need for further research.

In the RVD group, the proportion of patients with RVSP >50 mmHg was higher, and the QCG-PHTN score increased with increasing RVSP, showing good utility. However, there was a wider distribution of QCG-PHTN scores in the high RVSP group (especially >64 mmHg) compared to the relatively low RVSP group, which can be explained by the fact that RVSP is not an absolute reflection of pulmonary hypertension and is influenced by the hemodynamics of the RV. On echocardiography, RVSP is mainly calculated from the tricuspid regurgitation maximal velocity (TR Vmax) and the collapsibility of the inferior vena cava [5]. In patients with RV dysfunction, the TR Vmax is also reduced because the contractile force of the RV is reduced, which may cause this result.

This study has several limitations. Firstly, it is a single-center, retrospective study with a small sample size, which may limit the generalizability of findings. Therefore, a larger, multicenter, prospective study is needed. Second, we defined RV dysfunction based on echocardiography, which relies on the expertise of a trained examiner, and RVSP is not an absolute reflection of pulmonary hypertension as it is dependent on RV dysfunction and cardiac physiology, and therefore needs to reflect a more appropriate endpoint. Finally, since this study was based on patients with already diagnosed PE, a real-world point-of-care screening test would need to incorporate a variety of patient clinical conditions.

## Conclusion

In conclusion, when predicting RV dysfunction of digital ECG biomarker in patients with acute PE using ECGs, smartphone software can more accurately assess the presence or absence of RV dysfunction compared to traditional methods by clinical specialists. Particularly, the smartphone software demonstrates a high negative predictive value, suggesting the potential to omit or delay costly and time-consuming echocardiography in patients with a low risk.

## Data Availability

All data produced in the present study are available upon reasonable request to the authors

## Acknowledgement

This research was supported by a grant of the Korea Health Technology R&D Project through the Korea Health Industry Development Institute (KHIDI), funded by the Ministry of Health & Welfare, Republic of Korea (grant number: RS-2023-00265933)

## Author Contributions

Conceptualization: Yoo Jin Choi, Min Ji Park, Joonghee Kim, Moon Seung Soh Data curation: Yoo Jin Choi, Min Ji Park, Seo-Yoon Kim Formal analysis: Yoo Jin Choi, Min Ji Park, Moon Seung Soh Methodology: Youngjin Cho, Joonghee Kim, Eunkyoung Lee, Dahyeon Son Writing - original draft: Yoo Jin Choi, Min Ji Park Writing - review & editing: Youngjin Cho, Joonghee Kim, Seo-Yoon Kim, Moon Seung Soh

