## Supplementary figures and images for "Screening for Right Ventricular Dysfunction in the Emergency Department Using a Smartphone ECG Analysis Application: An External Validation Study with Acute Pulmonary Embolism Patients"

### Supplemental figure 1

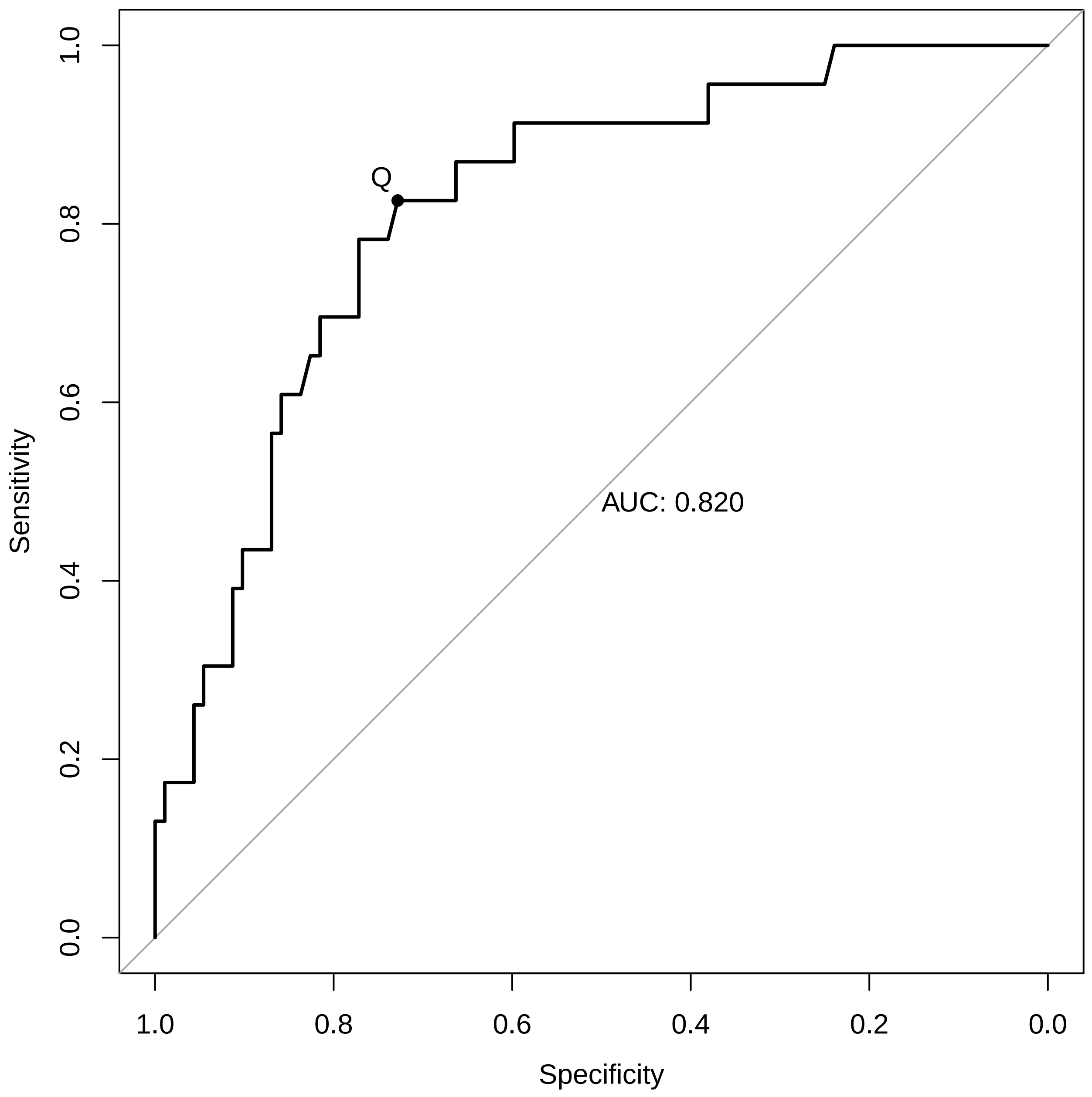

### Supplemental figure 2

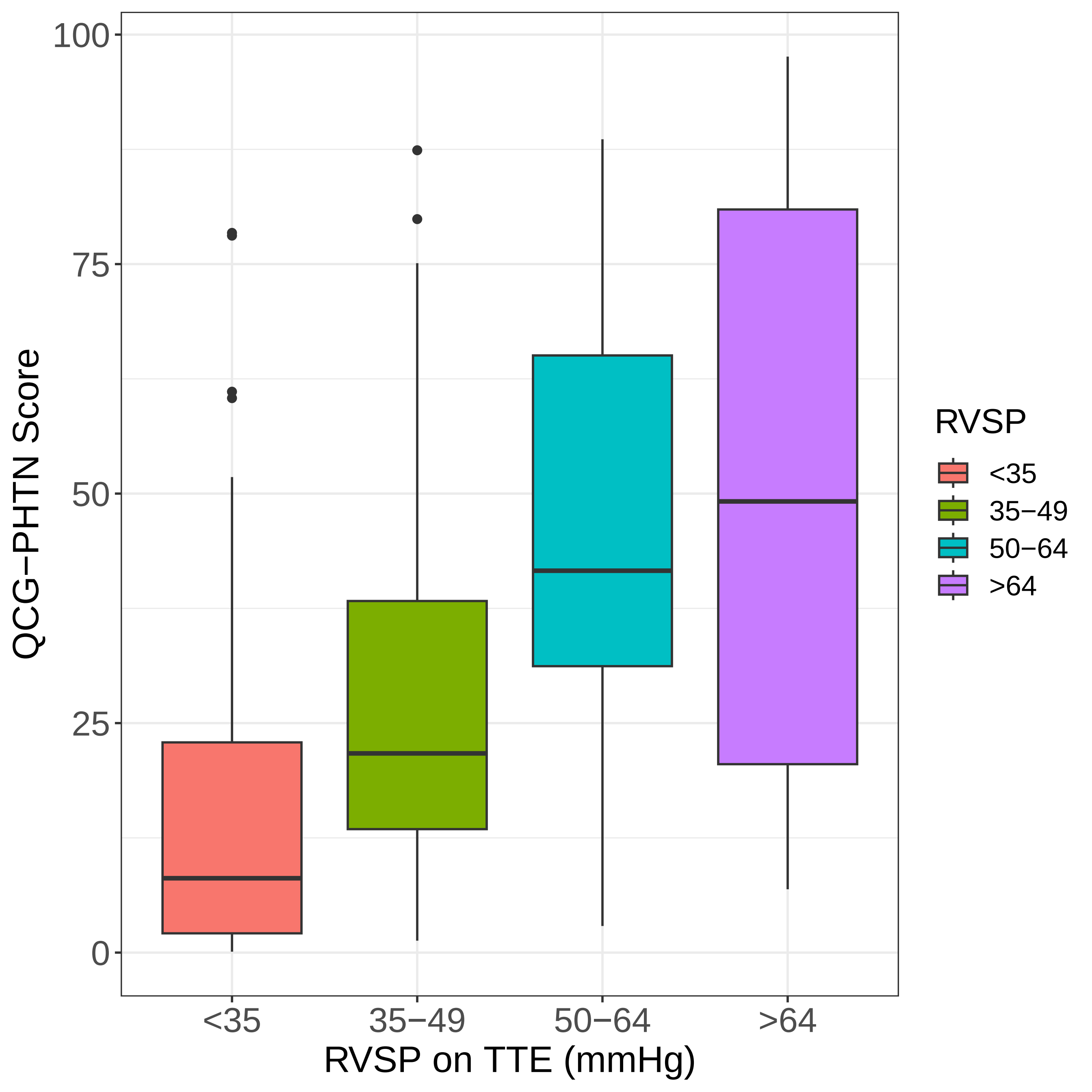
