## Supplemental figure legends for "Screening for Right Ventricular Dysfunction in the Emergency Department Using a Smartphone ECG Analysis Application: An External Validation Study with Acute Pulmonary Embolism Patients"

Supplementary Fig. 1. QCG-PHTN score across RVSP groups (RVSP, right ventricular systolic pressure)

Supplementary Fig. 2. Performance of QCG-PHTN on increased RVSP (RVSP >= 50mmHg)

Supplementary table 1. Performance of binarized QCG-RVDys and human experts on identifying RV dysfunction from ECG

| Biomarker or expert | AUC (95% CI) | P for difference | Sensitivity (95% CI) | Specificity (95% CI) | PPV (95% CI) | NPV (95% CI) |
| --- | --- | --- | --- | --- | --- | --- |
| QCG-RVDys (Binarized) | 0.845 (0.778-0.911) | - | 91.2 (82.4-100.0) | 77.8 (69.1-86.4) | 63.3 (54.4-73.9) | 95.5 (90.8-100.0) |
| Expert #1 | 0.676 (0.583-0.770) | 0.005 | 68.6 (51.4-82.9) | 66.7 (56.8-76.5) | 47.2 (38.2-57.2) | 83.1 (75.8-90.5) |
| Expert #2 | 0.659 (0.569-0.749) | <0.001 | 42.9 (28.6-60.0) | 88.9 (81.5-95.1) | 62.5 (45.5-81.0) | 78.2 (73.4-83.7) |
| Expert #3 | 0.628 (0.531-0.724) | <0.001 | 51.4 (34.3-68.6) | 74.1 (64.2-84.0) | 46.2 (34.2-60.0) | 77.9 (71.9-84.6) |
| Expert #4 | 0.683 (0.596-0.771) | 0.002 | 42.9 (28.6-60.0) | 93.8 (87.7-98.8) | 75.0 (57.7-92.9) | 79.2 (74.7-84.1) |
| Expert #5 | 0.658 (0.562-0.754) | 0.001 | 60.0 (45.7-77.1) | 71.6 (62.9-81.5) | 47.8 (37.5-60.0) | 80.6 (74.3-87.8) |

AUC, area under the curve; CI, confidence interval; PPV, positive predictive value; NPV, negative predictive value; QCG, quantitative electrocardiography; RVDys, right ventricular dysfunction;

Supplementary table 2. performance of binarized QCG-PHTN on increased RVSP (RVSP >= 50mmHg)

| AUC | Sensitivity | Specificity | PPV | NPV | Threshold |
| --- | --- | --- | --- | --- | --- |
| 0.820 (0.728-0.912) | 82.6 (65.2-95.7) | 72.8 (64.1-81.5) | 43.2 (34.7-54.1) | 94.4 (89.4-98.6) | 0.259 |

AUC, area under the curve; PPV, positive predictive value; NPV, negative predictive value;
